# Extending an Established Speech Recognition Threshold Procedure for Use in Noise: An Exploratory Study

**DOI:** 10.64898/2026.11.02.26366907

**Authors:** Josée Lagacé, Héloïse Lessard-Dostie

## Abstract

**Purpose:** This exploratory study examined whether an established clinical procedure for determining speech recognition thresholds in quiet, as outlined by the American Speech-Language-Hearing Association (1988), could be extended to a competing-noise condition using existing recorded speech materials. The objectives were to determine whether the resulting speech recognition thresholds in noise (SRTinNs) were broadly consistent with findings from established speech-in-noise measures and to examine their associations with pure-tone thresholds.

**Methods:** Twenty-five adults participated in this exploratory study: 15 with sensorineural hearing loss and 10 with normal audiograms. SRTinNs were estimated using an adaptation of the established threshold-seeking procedure with Canadian French monosyllabic words presented against continuous eight-talker babble. SRTinNs were compared between groups, and associations with pure-tone averages were examined among participants with hearing loss.

**Results:** Mean SRTinNs were +7.2 dB signal-to-noise ratio (SNR) for participants with hearing loss and +1.7 dB SNR for those with normal audiograms, representing a significant difference of 5.5 dB SNR (p = .02). Among participants with hearing loss, SRTinNs were significantly correlated with conventional pure-tone averages (r = .54, p = .04) and high-frequency pure-tone averages (r = .64, p = .01).

**Conclusions:** The findings provide preliminary support for further investigation of extending an established clinical threshold-seeking procedure to competing-noise conditions. Further investigation of this approach may help expand access to speech-in-noise assessment in linguistic contexts where suitable recorded speech materials are available but dedicated speech-in-noise tests are lacking.

## INTRODUCTION

Difficulty understanding speech in noisy everyday environments is among the most common complaints reported by adults seeking audiological care (Kochkin, 2010; Tremblay et al, 2015). Accordingly, speech-in-noise assessment has been recommended as an important component of a comprehensive audiological evaluation (Wilson, 2011; British Society of Audiology, 2019; Davidson et al, 2021; Fitzgerald et al, 2024).

Nevertheless, speech-in-noise assessment has yet to become a standard component of routine audiological practice, where speech recognition testing in quiet remains the primary functional measure (Parmar et al, 2022). Several practical barriers have been identified as contributing to the gap between the recognized clinical value of speech-in-noise assessment and its integration into routine audiological practice (Reynard et al, 2022; Fitzgerald et al, 2023; Billings et al, 2024). These include limited access to appropriate tests in the language of the patient (Reynard et al, 2022), additional administration time (Fitzgerald et al, 2023), and, for some procedures, requirements for specialized equipment (Reynard et al, 2022). The challenge is not to further establish the clinical value of speech-in-noise assessment, but to identify practical approaches that facilitate its integration into routine audiological care.

Some speech-in-noise tests are relatively simple to administer and can be implemented using equipment commonly available in audiology clinics. The Quick Speech-in-Noise test (QuickSIN; Killion et al., 2004) is one example and is among the commonly used speech-in-noise measures in English-speaking clinical settings (Parmar et al., 2022). However, similarly convenient and easy-to-administer speech-in-noise tests are not available across all languages. Access to linguistically appropriate speech-in-noise tests remains a challenge in some linguistic contexts (e.g., Lagacé & Lefebvre, 2017). When validated tests are not available in a patient’s primary language, clinicians have limited options for assessing speech-in-noise performance.

Developing and validating language-specific speech-in-noise tests is one approach to improving equitable access to speech-in-noise assessment. However, the development of such tests can require substantial time and resources. In some linguistic contexts, an alternative worth investigating may be to use existing recorded speech materials that have been developed and validated for speech audiometry in quiet (e.g., Trimmis et al., 2006; Nissen et al., 2011; Najem & Marie, 2021) to assess speech recognition in noise. When such materials are available and appropriately calibrated, rather than developing an entirely new speech-in-noise test, it may be possible to adapt an existing threshold-seeking procedure for use in noise. Such an approach would retain the principles of an established threshold-seeking procedure while introducing controlled competing noise during presentation of the speech stimuli.

One potential candidate is the procedure originally described by Tillman and Olsen (1973) for estimating speech recognition thresholds in quiet and subsequently incorporated into the American Speech-Language-Hearing Association (ASHA, 1988) guidelines. This procedure continues to be described in contemporary audiology textbooks as an approach for determining speech recognition thresholds in quiet (Katz et al., 2015; Gelfand & Calandruccio, 2023).

The present study examined whether the procedure outlined by ASHA (1988) for determining speech recognition thresholds in quiet could be extended to a competing-noise condition using validated recorded speech materials. The first objective of this exploratory study was to determine whether the resulting speech recognition threshold in noise (SRTinN) estimates in adults with and without hearing loss would be broadly consistent with values reported for comparable populations using established speech-in-noise measures. The second objective was to examine the association between SRTinN estimates and pure-tone audiometric thresholds.

## METHODS

### Recruitment

The present exploratory study was conducted in collaboration with three audiologists (HLD, SW, EGB) working at the Polyclinique de l’Oreille (Gatineau, Canada). The data collection protocol was approved by the University of Ottawa Research Ethics Board (#H 09-15-10). All participants provided written informed consent prior to participation.

Participants with hearing loss were recruited from the clientele attending the clinic. An invitation poster describing the study was displayed in the clinic waiting area, and the receptionist recorded the names of individuals who expressed interest in participating. Interested individuals were invited to participate in a testing session immediately following their clinical appointment. The research testing session involved only SRTinN measurement.

Participants in the normal-audiogram group were recruited through a general email announcement distributed by the program secretariat to staff and program collaborators. Individuals interested in participating contacted the research team directly. Participants in this group were not patients of the clinic where the study was conducted.

### Participants

To be included in the group of participants with hearing loss (hearing loss group), individuals were required to have sensorineural hearing loss, with air-conduction thresholds ≥25 dB HL at three or more frequencies between 0.25 and 8 kHz bilaterally. Individuals with a high-frequency pure-tone average (HFPTA; calculated from thresholds at 1, 2, and 4 kHz) greater than 55 dB HL in the better-hearing ear were excluded. This criterion was applied to reduce the likelihood that audibility of the 55-dB HL babble would be limited by the degree of hearing loss.

The hearing loss group included 15 adults (five women and ten men), aged 46 to 75 years (mean = 62.1 years, SD = 7.1 years). Individual pure-tone thresholds at each frequency in the better-hearing ear are reported in Table 1. The table also includes the conventional pure-tone average (PTA), calculated from thresholds at 0.5, 1, and 2 kHz, as well as the HFPTA.

**Table 1.** Individual pure-tone thresholds in the better-hearing ear, pure-tone averages, word-recognition scores in quiet, and speech recognition thresholds in noise (SRTinN) for participants with hearing loss, including group means and standard deviations (SD).

| Partici-pants | Age range (years) | Frequencies tested<br>Air conduction pure tone threshold (in dB HL) |  |  |  |  |  |  |  | PTA | HF PTA | Words in quiet | SRTinN (dB HL) | SRTinN (dB SNR) |
|---|---|---|---|---|---|---|---|---|---|---|---|---|---|---|
|  |  | 0.25 kHz | 0.5 kHz | 1 kHz | 2 kHz | 3 kHz | 4 kHz | 6 kHz | 8 kHz |  |  |  |  |  |
| HS-5 | 46-50 | 15 | 20 | 25 | 25 | 30 | 35 | 50 | 50 | 23 | 28 | 100 | 54 | -1 |
| HS-6 | 51-55 | 15 | 25 | 50 | 45 | 55 | 55 | 55 | 55 | 40 | 50 | 100 | 69 | 14 |
| HS-8 | 56-60 | 15 | 15 | 15 | 25 | 45 | 50 | 55 | 65 | 18 | 30 | 100 | 59 | 4 |
| HS-12 | 71-75 | 30 | 35 | 30 | 40 | 65 | 65 | 70 | 80 | 35 | 45 | 88 | 65 | 10 |
| SS-3 | 56-60 | 15 | 15 | 20 | 20 | 55 | 60 | 65 | 80 | 18 | 33 | 92 | 60 | 5 |
| SS-4 | 61-65 | 20 | 20 | 25 | 30 | 50 | 55 | 50 | 65 | 25 | 37 | 100 | 57 | 2 |
| SS-5 | 66-70 | 15 | 15 | 40 | 55 | 60 | 50 | 70 | 95 | 37 | 48 | 84 | 82 | 27 |
| SS-7 | 61-65 | 35 | 35 | 35 | 35 | 45 | 55 | 70 | 75 | 35 | 42 | 88 | 63 | 8 |
| SS-8 | 61-65 | 35 | 30 | 35 | 35 | 50 | 45 | 50 | 55 | 33 | 38 | 92 | 63 | 8 |
| SS-9 | 66-70 | 10 | 25 | 30 | 30 | 30 | 40 | 65 | 65 | 28 | 33 | 96 | 67 | 12 |
| SS-10 | 56-60 | 15 | 20 | 45 | 45 | 50 | 35 | 30 | 35 | 37 | 42 | 92 | 54 | -1 |
| ES-1 | 56-60 | 35 | 35 | 40 | 45 | 50 | 60 | NT | 65 | 40 | 48 | 92 | 66 | 11 |
| ES-3 | 61-65 | 20 | 20 | 25 | 45 | 55 | 50 | NT | 60 | 30 | 40 | 100 | 59 | 4 |
| ES-6 | 66-70 | 15 | 15 | 20 | 30 | 35 | 35 | 40 | 55 | 22 | 28 | 100 | 55 | 0 |
| ES-7 | 61-65 | 10 | 10 | 15 | 40 | 40 | 25 | 20 | 5 | 22 | 27 | 96 | 60 | 5 |
| MEAN<br>(SD) | 62<br>(7.1) | 20<br>(9.1) | 22.3<br>(8.2) | 30<br>(10.7) | 36.3<br>(9.7) | 47.7<br>(10.3) | 47.7<br>(11.5) | 53.1<br>(15.8) | 60.3<br>(21) | 29.6<br>(7.7) | 38<br>(7.8) | 95<br>(5.4) | 62.2<br>(7.2) | 7.2 |

Note: PTA = pure-tone average at 0.5, 1, and 2 kHz; HFPTA = high-frequency pure-tone average at 1, 2, and 4 kHz; SRTinN = speech recognition threshold in noise; SNR = signal-to-noise ratio; NT = not tested. Word-recognition scores in quiet are expressed as percentages. Pure-tone thresholds, PTA, HFPTA, and SRTinN are expressed in dB HL unless otherwise indicated.

As summarized in Table 1, participants had, on average, a sloping hearing loss, with mean thresholds ranging from the mild hearing loss range at 1 kHz to the moderately severe hearing loss range at 8 kHz in their better-hearing ear. Mean HFPTA was 38 dB HL (SD = 7.8 dB), with values ranging from 27 to 50 dB HL. Mean word-recognition performance in quiet was 95% (range: 84–100%).

To be included in the group of participants with normal audiograms (normal audiogram group), individuals were required to have air-conduction thresholds ≤20 dB HL at all tested frequencies (0.25, 0.5, 1, 2, 4, and 8 kHz) bilaterally. The normal audiogram group included 10 adults (six women and four men), aged 47 to 60 years (mean = 54.8 years, SD = 4.5 years). Individual pure-tone thresholds at each frequency in the better-hearing ear are reported in Table 2, along with the conventional PTA and HFPTA.

**Table 2.** Individual pure-tone thresholds in the better-hearing ear, pure-tone averages, word-recognition scores in quiet, and speech recognition thresholds in noise (SRTinN) for participants with normal audiograms. Group means and standard deviations (SD) are also reported.

| Parti-cipants | Age range (years) | Frequencies tested<br>Air conduction pure tone threshold (in dB HL) |  |  |  |  |  |  |  | PTA | HF PTA | Words in quiet | SRTinN (dB HL) | SRTinN (dB SNR) |
|---|---|---|---|---|---|---|---|---|---|---|---|---|---|---|
|  |  | 0.25 kHz | 0.5 kHz | 1 kHz | 2 kHz | 3 kHz | 4 kHz | 6 kHz | 8 kHz |  |  |  |  |  |
| EN-1 | 46-50 | 10 | 5 | 10 | 5 | NT | 0 | NT | 5 | 7 | 5 | 100 | 53 | 2 |
| EN-2 | 51-55 | 15 | 15 | 15 | 10 | NT | 5 | NT | 15 | 13 | 10 | 100 | 57 | 2 |
| EN-3 | 56-60 | 5 | 5 | 10 | 5 | NT | 10 | NT | 10 | 7 | 8 | 100 | 58 | 3 |
| EN-5 | 56-60 | 10 | 5 | 5 | 5 | NT | 5 | NT | 10 | 5 | 5 | 100 | 58 | 3 |
| EN-6 | 51-55 | 10 | 5 | 5 | 15 | NT | 5 | NT | 10 | 8 | 8 | 100 | 54 | -1 |
| EN-7 | 56-60 | 10 | 5 | 15 | 10 | NT | 20 | NT | 15 | 10 | 15 | 100 | 58 | 3 |
| EN-8 | 51-55 | 15 | 5 | 10 | 5 | NT | 15 | NT | 15 | 7 | 10 | 92 | 55 | 0 |
| EN-9 | 46-50 | 0 | 5 | 5 | 5 | NT | 5 | NT | 10 | 5 | 5 | 100 | 58 | 3 |
| EN-10 | 56-60 | 10 | 5 | 15 | 15 | NT | 10 | NT | 10 | 12 | 13 | 100 | 57 | 2 |
| EN-11 | 51-55 | 15 | 10 | 5 | 10 | NT | 5 | NT | 5 | 8 | 7 | 100 | 59 | 4 |
| MEAN (SD) | 55 (4.5) | 10 (4.7) | 6.5 (3.4) | 9.5 (4.4) | 8.5 (4.1) |  | 8 (5.9) |  | 10.5 (4) | 8 (3) | 9 (3) | 99 (3) | 56.7 (2) | 1.7 |

Note: PTA = pure-tone average at 0.5, 1, and 2 kHz; HFPTA = high-frequency pure-tone average at 1, 2, and 4 kHz; SRTinN = speech recognition threshold in noise; SNR = signal-to-noise ratio; NT = not tested. Word-recognition scores in quiet are expressed as percentages. Pure-tone thresholds, PTA, HFPTA, and SRTinN are expressed in dB HL unless otherwise indicated.

The mean HFPTA of the normal audiogram group was 9 dB HL (SD = 3 dB), with values ranging from 5 to 15 dB HL. Mean word-recognition performance in quiet was 99.2% (range: 92–100%).

All 25 participants reported no history of developmental language or learning difficulties or of cognitive, neurological, or attention disorders. For all participants, Canadian French was the first language learned and the language used in everyday conversation at least 50% of the time.

### Materials

For the present study, recordings from the *Test de Mots dans le Bruit* (TMB; Lagacé, 2010) were used to measure SRTinN. Presentation levels were controlled using an Interacoustics AC40 audiometer connected to the sound-file player. The recordings were presented through insert earphones.

The TMB consists of four lists of monosyllabic words, with the words and competing babble recorded on separate channels to allow their presentation levels to be controlled independently (Lagacé, 2010). The words were recorded by a female speaker of Canadian French, and word familiarity was established for children aged 6 years and older. The level of each recorded word was normalized to within ±2 dB of the root-mean-square level (69.3 dB SPL) of the complete word corpus (Lagacé, 2010). The competing noise consisted of an eight-talker French babble developed by Perrin and Grimault (2005).

Unlike recorded speech materials developed specifically for speech audiometry in quiet, the TMB includes a competing-noise recording. However, because the speech and babble were recorded on separate channels, their presentation levels could be controlled independently, including presentation of the speech without the babble. This feature made the TMB recordings suitable for examining whether the threshold-seeking procedure investigated in the present study could be applied under a controlled competing-noise condition.

### Procedure

Pure-tone thresholds and word-recognition performance in quiet were obtained on the same day as SRTinN testing by the same audiologists and using the same clinical procedures for both groups. Pure-tone air-conduction thresholds were measured according to ANSI (2019) and ASHA (2005) guidelines. For participants with hearing loss, these measures were part of the clinical assessment conducted that day, and the results were extracted from their clinical files. For participants of the normal audiogram group, these measures were obtained as part of the study immediately prior to SRTinN testing.

SRTinN was measured monaurally in the better-hearing ear, defined as the ear with the lower PTA (0.5, 1, and 2 kHz); when PTA was identical between ears, the right ear was tested. Throughout the procedure, the TMB babble was presented continuously and ipsilaterally at 55 dB HL, while the presentation level of the recorded words was varied according to the threshold-seeking procedure described below.

The protocol used in the present study differed from the procedure outlined by ASHA (1988) in two respects: (1) monosyllabic words were used instead of spondees, and (2) continuous ipsilateral babble was presented during testing rather than speech being presented in quiet. The first stage was used to determine the initial level. The first word from the assigned TMB list was presented at 30 dB SL relative to the PTA of the test ear. Participants were instructed to repeat the word presented after the warning beep and to guess when unsure. If the first word was repeated correctly, a second word was presented at the same level. The presentation level was then decreased in 10-dB steps, with two words presented at each level. Correctly repeating at least one of the two words was sufficient to proceed to the next 10-dB decrement. This descending procedure continued until neither of the two words presented at a given level was correctly repeated. This level was defined as the initial level. If neither of the first two words presented at 30 dB SL was correctly repeated, the procedure was restarted at 40 dB SL.

Once the initial level had been established, the starting level for threshold determination was set at 10 dB above the initial level. Using subsequent words from the same TMB list, two words were presented at the starting level, followed by successive pairs of words presented in 2-dB descending steps. Correct responses were accumulated across presentation levels. Consistent with the ASHA (1988) procedure, at least five of the first six words presented from the starting level had to be correctly repeated. If this criterion was not met, the first stage was repeated to establish a higher initial level, from which a new starting level was derived.

Testing continued in 2-dB descending steps, with two words presented at each level, until at least five of the final six words were incorrectly repeated or not repeated. SRTinN was then calculated by subtracting the total number of correctly repeated words from the starting level and adding the 1-dB correction factor specified in the ASHA (1988) procedure. Because the competing babble remained fixed at 55 dB HL throughout testing, the resulting threshold could also be expressed as an SNR by subtracting 55 dB from the SRTinN expressed in dB HL. A step-by-step example of the procedure is provided in Table 3.

**Table 3.** Procedure used to determine the speech recognition threshold in noise (SRTinN), illustrated with a fictitious case with a PTA of 45 dB HL.

| Stage | Presentation level (dB HL) | Responses | Decision / procedure |
|---|---|---|---|
| *Initial level determination* | 75 | ✓✓ | Initial presentation level = 30 dB SL re: PTA (0.5, 1, 2 kHz) |
|  | 65 | ✓× | Decrease by 10 dB |
|  | 55 | ×× | Both words missed; initial level = 55 dB HL |
| *Starting level* | 65 | ✓✓ | *Starting level* = 10 dB above the initial level |
|  | 63 | ✓✓ | Decrease in 2-dB steps |
|  | 61 | ✓✓ | At least 5 of the first 6 words correctly repeated |
|  | 59 | ✓× | Continue in 2-dB steps |
|  | 57 | ×× |  |
|  | 55 | ✓× |  |
|  | 53 | × ✓ |  |
|  | 51 | ×× |  |
|  | 49 | ×× | At least 5 of the last 6 words missed |
| *SRTinN = starting level − total number of correctly repeated words + 1 dB*<br>*= 65 − 9 + 1 = 57 dB HL (+2 dB SNR)* |  |  |  |

Note: ✓ = correctly repeated word; × = missed or incorrectly repeated word. The eight-talker babble from the TMB recording was presented continuously and ipsilaterally at 55 dB HL throughout the procedure.

A different TMB list was assigned across participants according to a Latin-square sequence. When completion of the procedure required more words than were available in the assigned list, testing continued with the next sequential list. For example, when testing began with List 2, List 3 was used if additional words were required.

### Statistical analyses

SRTinN values were compared between the hearing-loss and normal-audiogram groups using a Mann–Whitney *U* test. Among participants with hearing loss, Pearson correlation analyses were conducted to examine the associations between SRTinN and conventional PTA and between SRTinN and HFPTA. Statistical significance was set at α = .05. Analyses were performed using Microsoft Excel, with the Mann–Whitney *U* test calculated from ranked data using a two-tailed normal approximation corrected for ties and continuity.

## RESULTS

### SRTinN estimates

SRTinN estimates for individual participants are presented in Tables 1 and 2. Participants with hearing loss had a mean SRTinN of 62.2 dB HL (SD = 7.2 dB), corresponding to +7.2 dB SNR. Participants with normal audiograms had a mean SRTinN of 56.7 dB HL (SD = 2.0 dB), corresponding to +1.7 dB SNR. Thus, mean SRTinN differed by 5.5 dB SNR between the two groups. A Mann–Whitney U test indicated a significant difference in SRTinN between participants with hearing loss and those with normal audiograms (*U* = 31.5, *p* = 0.02).

### Association between SRTinN and pure-tone thresholds

Among participants with hearing loss, SRTinN was significantly correlated with conventional PTA (*r* = 0.54, *p* = 0.04), corresponding to 29% shared variance (*R*² = 0.29). A descriptively stronger association was observed between SRTinN and HFPTA (*r* = 0.64, *p* = 0.01), corresponding to 41% shared variance (*R*² = 0.41).

## DISCUSSION

The present exploratory study examined whether an established clinical procedure for determining speech recognition thresholds in quiet, as outlined by ASHA (1988), could be extended to a competing-noise condition. The main objective was to determine whether the resulting SRTinN estimates would be consistent with values reported for comparable populations using established speech-in-noise measures. Participants with hearing loss had a mean SRTinN of +7.2 dB SNR, compared with +1.7 dB SNR for participants with normal audiograms, corresponding to a mean difference of 5.5 dB SNR. This overall pattern is broadly consistent with earlier reports using other speech-in-noise tests or procedures, in which listeners with hearing loss generally required more favorable SNRs than listeners with normal audiograms (Killion et al., 2004; Wilson, 2011; Fitzgerald et al., 2023; Billings et al., 2024). In terms of the magnitude of the difference between group means, Wilson et al. (2003), using the Words-in-Noise (WIN) test, reported an approximately 5-dB difference between young adults with normal audiograms and older adults with mild hearing loss, with mean SRTinNs of approximately +4 and +9 dB SNR, respectively. Although the magnitude of this difference is like that observed in the present study, the groups in Wilson et al. differed in both hearing status and age, precluding attribution of the group difference to hearing loss alone.

Group mean SRTinNs, however, do not fully capture variability in individual performance. As reported in previous studies, greater variability in speech-in-noise performance was observed among participants with hearing loss than among those with normal audiograms (Killion et al., 2004; Wilson, 2011; Fitzgerald et al., 2023; Billings et al., 2024). In the present study, SRTinNs ranged from −1 to +27 dB SNR among participants with hearing loss, compared with −2 to +4 dB SNR among participants with normal audiograms. Notably, two participants with hearing loss obtained SRTinNs of −1 dB SNR, placing their performance within the range observed among participants with normal audiograms, whereas other participants with hearing loss required substantially more favorable SNRs. Thus, although hearing loss was associated with poorer SRTinN at the group level, the wide range of individual values indicates that hearing status alone did not fully account for speech-in-noise performance.

The second objective of the study was to examine the association between SRTinN estimates and pure-tone audiometric thresholds. Among participants with hearing loss, SRTinN was significantly associated with pure-tone hearing sensitivity, although the associations accounted for only part of the observed variance. The shared variance between SRTinN and conventional PTA was 29%, whereas the shared variance between SRTinN and HFPTA was 41%. This pattern is consistent with previous reports showing an association between high-frequency hearing sensitivity and speech-in-noise performance (e.g., Wilson, 2011). Nevertheless, substantial variance in SRTinN remained unaccounted for even when HFPTA was considered, indicating that pure-tone hearing sensitivity and speech-in-noise performance provide related but nonredundant information. This finding is consistent with the longstanding recommendation that hearing sensitivity and speech recognition in competing noise be assessed separately to provide a more complete characterization of an individual’s auditory abilities (Killion, 2002; Wilson, 2011; Fitzgerald et al., 2023; Billings et al., 2024).

A similar distinction is observed when speech-in-noise performance is considered in relation to word recognition in quiet. Despite high mean word-recognition scores in quiet in both groups (95% for participants with hearing loss and 99% for those with normal audiograms), SRTinN differed between the groups and showed substantial variability among participants with hearing loss. Thus, in the present sample, performance in quiet did not reflect the range of performance observed under competing-noise conditions. This observation is consistent with previous reports indicating that speech recognition in quiet and speech recognition in noise provide different and complementary information about auditory performance (Wilson, 2011; Fitzgerald et al, 2023).

The findings of this exploratory study should be interpreted in light of its limitations. First, the relatively small sample size and recruitment of adults from a single hearing health clinic limit the generalizability of the findings. The present results therefore cannot be assumed to extend to other age groups or to populations with different hearing characteristics. In addition, because the procedure was evaluated using Canadian French speech materials, the SRTinN values obtained in this study cannot be assumed to generalize to speech materials in other languages. Second, test–retest reliability was not assessed; therefore, the stability and reproducibility of SRTinN estimates obtained with the adapted procedure remain to be established.

The present findings provide preliminary support for further investigation of extending an established clinical threshold-seeking procedure to a competing-noise condition using existing recorded speech materials. Importantly, although validated speech-in-noise tests are not available in all languages, recorded materials developed for conventional speech audiometry in quiet are available in a number of languages (Trimmis et al., 2006; Nissen et al., 2011; Najem and Marie, 2021). These materials could provide a starting point for evaluating the adapted procedure in other linguistic contexts, rather than requiring the development of an entirely new language-specific speech-in-noise test. Such applications would require linguistically appropriate speech materials, adequate evaluation of the relevant acoustic characteristics and calibration of both the speech and competing-noise materials, and further investigation of the psychometric properties of the adapted procedure, including its test–retest reliability. If these requirements can be met, the proposed approach may help reduce one of the practical barriers to incorporating speech-in-noise assessment into routine audiological care and ultimately contribute to more equitable access to such assessment across a wider range of linguistic contexts.

## Data Availability

All data produced in the present work are contained in the manuscript.

## Acknowledgements

This study received no external funding. The authors would like to thank the audiologists who contributed to data collection, Sophie Waridel and Éliane Gauvin-Bourdages, as well as the Polyclinique de l’oreille for allowing the study to be conducted in its clinics. The authors are grateful to all participants for the time they voluntarily devoted to this project. They also wish to thank Nisrine Zamjaoui, Benoît Jutras, and Catherine Belzile for their contributions to earlier versions of the manuscript.

## Declaration of Generative AI Use

OpenAI’s ChatGPT was used during manuscript preparation as an editorial support tool to assist with language and clarity. The authors take full responsibility for the accuracy and integrity of the final manuscript.

## LIST OF ABBREVIATIONS AND ACRONYMS

HFPTA: high-frequency pure-tone average PTA - pure tone average
SRTinN: speech recognition threshold in noise
TMB: *Test de Mots dans le Bruit*

